# Impact of the Mindfulness-Based Blood Pressure Reduction (MB-BP) Program on Cardiovascular Health: A Randomized Clinical Trial

**DOI:** 10.1101/2025.05.27.25328464

**Authors:** Fan Wu, LaPrincess C. Brewer, Vinicius V. Neves, Matthew M. Scarpaci, Jeffrey A. Proulx, Eric B. Loucks

## Abstract

**Background:** Mindfulness-based interventions may improve cardiovascular health (CVH) by supporting behavioral change across multiple risk factors. This study evaluated the impact of Mindfulness-Based Blood Pressure Reduction (MB-BP), a mindfulness program targeting hypertension-related behaviors, on CVH using the American Heart Association’s Life’s Essential 8 framework.

**Methods:** This secondary analysis of a preregistered, parallel-group, phase 2 randomized clinical trial evaluated the effects of MB-BP on CVH in 201 participants with elevated office BP (≥120/80 mmHg). The MB-BP group (n=101) received an 8-week program focused on mindfulness training and education targeting diet, physical activity, medication adherence, alcohol use, and stress, whereas the control group (n=100) received enhanced usual care. CVH was assessed using available Life’s Essential 8 components: systolic blood pressure, body mass index (BMI), diet (DASH adherence), physical activity, smoking, and sleep duration. Generalized estimating equations evaluated intervention effects through six months.

**Results:** At 6 months follow-up, MB-BP participants significantly improved composite CVH scores compared to controls (standardized mean difference: 0.144; 95% CI: 0.023–0.266). Non-significant improvements were observed across most CVH components in MB-BP vs. control, including systolic blood pressure (−4.95 mmHg), DASH diet score (+0.27), physical activity (+47.9 MET-min/week), sleep duration (+0.34 hours/night), and BMI (−0.28 kg/m²). No significant changes were observed for smoking, likely due to the low baseline prevalence.

**Conclusions:** MB-BP led to modest but clinically significant improvements in CVH, driven by multiple Life’s Essential 8 components. These findings suggest that MB-BP may be an effective behavioral intervention to support CVH and reduce risk for cardiovascular disease.

**ClinicalTrials.gov Preregistration Identifiers:** NCT03256890, NCT03859076

---

Cardiovascular disease (CVD) remains the leading cause of mortality worldwide, responsible for an estimated 19.4 million deaths in 2021.^1^ In response, public health initiatives have expanded their focus from treating CVD alone to promoting overall cardiovascular health (CVH), aiming to enhance quality of life and reduce long-term disease risk.^2^ The American Heart Association defines CVH through eight core metrics—physical activity, diet, body mass index (BMI), smoking status, sleep, blood lipids, blood glucose, and blood pressure—collectively known as *Life’s Essential 8*.^3^

Of the top 20 risk factors contributing to years of life lost in the United States, 11 are directly related to components of CVH. In 2021, the four leading contributors were high systolic blood pressure, tobacco use, high fasting plasma glucose, and elevated BMI.^4^ National Health and Nutrition Examination Survey (NHANES) data from 1990 to 2021 indicate that while some aspects of CVH have improved—such as reduced smoking prevalence, lower systolic blood pressure, and increased consumption of whole grains—others have worsened. Specifically, there has been a rise in fasting glucose levels, BMI, and LDL cholesterol, along with decreased intake of fruits and vegetables and increased intake of processed meat and sodium.^4^

In recognition of the multifaceted drivers of CVH, the American Heart Association’s Presidential Advisory on Life’s Essential 8 emphasized the importance of psychological well-being and social determinants of health, identifying mindfulness as a promising influence on CVH.^5^

Transdiagnostic interventions—those that address multiple coexisting conditions, such as hypertension, obesity, hyperlipidemia, and diabetes—offer a potentially efficient means of improving population-level CVH while reducing CVD risk.^6,7^ One class of transdiagnostic interventions showing promise is mindfulness-based programs. Mindfulness has been described as comprising three key elements: (1) present-moment awareness of thoughts, emotions, and physical sensations; (2) the quality of that awareness, which includes non-judgment, curiosity, compassion, and discernment; and (3) remembering, such as remembering to bring one’s wisdom to the present moment and apply it skillfully.^8–10^

From a public health perspective, mindfulness has the potential to enhance intrinsic motivation, improve engagement, and make health interventions more meaningful—factors shown to improve adherence and long-term outcomes.^11,12^ Moreover, mindfulness may reduce stress and emotional reactivity that often accompany behavior change efforts, thereby lowering psychological barriers to sustained engagement.^12^ These qualities make mindfulness a compelling tool not only for clinical outcomes but also for supporting long-term behavior change in public health settings.

To conceptualize how mindfulness training might influence CVH, a theoretical framework was developed that emphasizes the cultivation of mindfulness skills—such as self-awareness, attention control, and emotion regulation—and their application to behaviors and risk factors relevant to CVD prevention (e.g., diet, physical activity, sleep, BMI, blood pressure, cholesterol).^12^ This framework is summarized in **Figure 1**.

**Figure 1.**
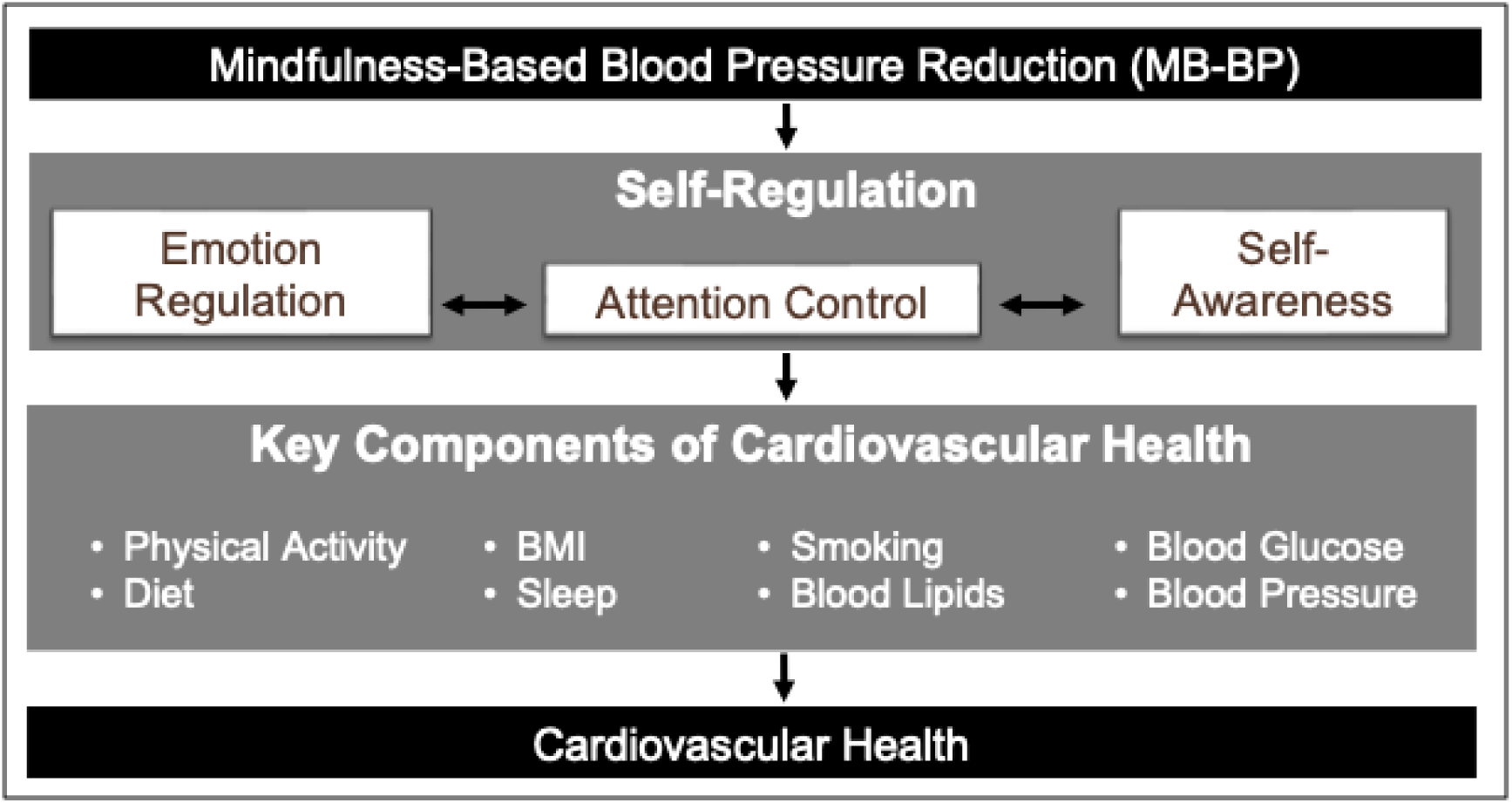
Theoretical framework for Mindfulness-Based Blood Pressure Reduction (MB-BP). MB-BP is theorized to enhance self-regulation processes (emotion regulation, attention control, and self-awareness), which influence key cardiovascular health components, including physical activity, diet, sleep, BMI, smoking, blood glucose, blood lipids, and blood pressure, ultimately improving overall cardiovascular health.

Emerging evidence suggests that mindfulness training may positively impact individual components of CVH, including dietary behavior, physical activity, blood pressure, weight management, and sleep quality.^13–16^ However, there is a paucity of research examining mindfulness interventions that target multiple CVH components simultaneously. To address this gap, the *Mindfulness-Based Blood Pressure Reduction (MB-BP)* program was developed to apply mindfulness skills to the behavioral and psychosocial drivers of hypertension, including Life’s Essential 8 components such as diet, physical activity, and weight regulation.^17^

While an observational study involving 382 participants demonstrated a cross-sectional association between dispositional mindfulness and cardiovascular health,^18^ there remains limited randomized trial evidence evaluating the impact of mindfulness training on CVH outcomes. Therefore, the objective of this study was to evaluate the effects of MB-BP on overall CVH at 6-month follow-up using a randomized controlled trial design.

## Methods

### Study Sample and Design

The MB-BP Study was a phase 2, parallel-group randomized clinical trial assessing the efficacy of group-based mindfulness intervention. This study focused on addressing determinants of blood pressure, many that are components of Life’s Essential 8 (e.g., diet, PA, weight loss) alongside other blood pressure drivers such as stress reactivity, alcohol consumption, and antihypertensive medication adherence.

Participant recruitment and assessments took place from June 2017 to November 2020, using community advertisements and referrals from healthcare and public health providers.^19^ The study was registered on ClinicalTrials.gov at the onset of participant recruitment (applied in July 2017, approved in August 2017) and prior to outcome assessment in participants. Data were not examined prior to registration. This study followed CONSORT guidelines for RCT reporting.^20^

Participants included adults aged 18 years or older who had elevated blood pressure (SBP ≥120 mm Hg or DBP ≥80 mm Hg) in both of two in-person assessments at least one week apart at baseline.^13^ The study excluded those who meditated more than once a week, were unable to communicate in English, had severe medical conditions that precluded regular class attendance, had substance use or eating disorder, had suicidal ideation, history of bipolar disorder, psychotic disorder, or self-injurious behavior. The Brown University institutional review board approved this study protocol on June 12, 2017 (protocol number 1412001171). All participants provided written informed consent.

Data for this study were meta-analyzed from two pre-registered clinical trials that utilized identical methodologies. Both were part of a larger project funded by a National Institutes of Health UH2/UH3 cooperative agreement. The primary outcomes were pre-registered in clinicalTrials.gov (#NCT03256890, #NCT03256890) and reported elsewhere.^13,14^

### Blinding and Randomization

Those participants who passed the screening and assessment performed at baseline were randomized in a 1:1 ratio either to participate in the MB-BP group or control group.

Randomization was stratified by 2 potential determinants of blood pressure at follow-up: sex and BP status (i.e., SBP≥140 mm Hg or DBP ≥90 mm Hg vs other). Randomization took place approximately a week before the start of the MB-BP classification period, and participants were notified of group assignments within 24 hours of randomization.^17^ One of the researchers blinded to the participant identity performed the randomization using Research Randomizer Version 4.0 software. The senior project manager communicated the group assignments to the participants. Staff conducting follow-up activities were blinded to group allocation, with the exception of events relatedness inquiry which was completed by a separate staff member.

### Intervention Descriptions and Theoretical Framework

The MB-BP program was adapted primarily from the Mindfulness-Based Stress Reduction (MBSR) program, but also included elements inspired by Mindfulness-Based Cognitive Therapy, and Acceptance and Commitment Therapy.^21^ Participants were encouraged to practice mindfulness at home for at least 45 minutes per day, six days a week after finishing group orientation sessions. The unique areas of MB-BP, described in detail in supplementary material **Table S1** and elsewhere^13,17^ were education about hypertension risk factors, along with specific mindfulness modules focused on mindful awareness of diet, physical activity, medication adherence, alcohol consumption, stress, and social support for behavior change. MB-BP builds a foundation of mindfulness skills (e.g., meditation, yoga, self-awareness, attention control, emotion regulation) through the MBSR curriculum. The program then directs those skills towards participants’ adhering to behaviors that can lower BP (**Figure 1**).^17^ MB-BP participants have their BP and hypertension risk factors directly assessed at baseline, and are provided with this information along with the recommended best practices and national guideline-based standards for blood pressure control. MB-BP encourages participants to explore personal readiness for change in the different hypertension self-management behaviors, and trains participants to utilize mindfulness practices to engage with those behaviors that they choose.

MB-BP was led by qualified MBSR instructors with expertise in cardiovascular disease etiology, treatment, and prevention. They were further trained and certified to teach MB-BP. Classes were provided in-person for the first ten cohorts (n=173), while the final two cohorts (n=28) began in-person and shifted to an online format via Zoom because of the COVID-19 pandemic. These classes were held in Providence, Rhode Island at the Brown University School of Public Health building, and at a community health center serving a low-income area.

Participants randomized to the intervention were provided with a home blood pressure (BP) monitor (Omron, model BP786N) and standardized instructions. Participants’ primary care physicians were contacted if their office blood pressure was ≥140/90 mmHg or were offered assistance in finding a primary care physician if they did not already have one. Participants randomized to the enhanced usual care arm received the same home BP device, training process, and similar primary care options, but without mindfulness training. Participants in the control group received an educational brochure on high blood pressure from the American Heart Association (product code 50-1731). In this study, treatment fidelity strategies were applied in the domains of study design, provider training, treatment delivery, receipt of treatment, and enactment of treatment skills, using the recommended strategies by the National Institutes of Health Behavior Change Consortium,^22^ described elsewhere.^17^ The instructors were evaluated for intervention fidelity and were required to maintain over 90% adherence to the curriculum.

### Outcomes

The primary outcome was CVH assessed at baseline, three months, and six months using six components of the American Heart Association’s Life’s Essential 8: blood pressure, BMI, smoking status, sleep duration, physical activity, and dietary pattern. Components were scored according to Life’s Essential 8 guidelines,^5^ which assign 0–100 points per component based on categorical thresholds (e.g., BMI <25 = 100; 25.0–29.9 = 70; 30.0–34.9 = 30; 35.0–39.9 = 15; ≥40 = 0). The overall CVH score was calculated as the mean of available components, yielding a score from 0 to 100, with higher scores indicating better CVH. Because the scoring system uses broad categories that may not detect meaningful within-category changes (e.g., a 31-lb range within BMI 25.0–29.9 for someone who is 5’8” tall), we prioritized analyses using standardized (z-scored) means for each component, while reporting both scoring approaches. As the MB-BP study did not collect blood samples, glucose and lipid levels were not included; results should be interpreted with this limitation in mind. Methods for assessing each CVH component are detailed below.

*Blood pressure* assessment occurred in a dedicated room at the Brown University School of Public Health using a calibrated Omron HEM-705CPN automated blood pressure monitor. The mean of the second and third blood pressure readings at each assessment was used for analyses, following best practices.^23^

*BMI* was determined based on weight and height measured with participants in light clothing without shoes, using a calibrated stadiometer (SECA, Hamburg, Germany) and weighing scale (SECA, Model 22089, Hamburg, Germany).

*Current cigarette smoking* was assessed via self-report of number of cigarettes smoked per day.

*Sleep duration* (average hours of sleep per night during the past month) was assessed using the Pittsburgh Sleep Quality Index.^24,25^

*Physical activity* during the prior week was measured using the International Physical Activity Questionnaire as total MET (metabolic equivalent of task)-minutes per week of physical activity.^26^

*Dietary Approaches to Stop Hypertension (DASH)-consistent diet* was assessed using the Harvard 163-item 2007 Grid Food Frequency Questionnaire,^27^ and coding of DASH diet adherence using methods developed by Folsom et al.^28^

### Other Relevant Baseline Non-CVH Variables

Alongside self-reported age, race and education other modifiable variables were provided in the baseline table to allow readers to evaluate characteristics of participants in the MB-BP and control groups at baseline. Effects on these variables have been reported elsewhere.^13,14^

*Antihypertensive medication* usage was collected directly from subjects’ medicine bottles at each assessment (baseline, 3 months follow-up and 6 months follow-up), recording prescribed medication type, dose and frequency.

*Alcohol consumption* was assessed using a modified Centers for Disease Control and Prevention Behavioral Factor Surveillance System Questionnaire, showing concurrent validity with other nationally-representative survey measures.^29^

*Sedentary behavior* during the prior week was measured using the International Physical Activity Questionnaire as time spent in sedentary activities.^26^

*Stress* levels were calculated using the validated 14-item Perceived Stress Scale (scale range: 0-56).^30^

*Mindfulness* levels were calculated with the validated Five-Facet Mindfulness Questionnaire (scale range: 39-195).^31^

Systematic monitoring for adverse events was undertaken with a particular view on clinically significant changes in anxiety and depression symptoms. Monthly online safety checks monitor serious adverse events and physical injuries. Any adverse events were recorded by the data safety monitoring plan, with reporting to the principal investigator and the chair of the data safety monitoring board.

### Statistical Analysis

We analyzed the effect of the MB-BP program compared with the control condition at both 3- and 6-months using generalized estimating equations (GEE) with an identity link and autoregressive covariance structure.^32^ CVH was evaluated as a variable ranging from a score of 0-100 using methods consistent with Lloyd-Jones et al described in the Outcomes section above,^5^ and as a continuous composite score using Z-scores to take advantage of the continuous nature of the outcome variables, the latter approach which should enhance sensitivity and statistical power. Unadjusted analyses were performed to avoid possible bias and imprecision associated with covariate adjustment, particularly when adjusting for baseline measures of the primary outcome.^33–35^ Analyses adjusted for the strata used to randomize participants (i.e., sex and BP status) can increase statistical power.^36^ Marginal means were reported as the estimate for each model. We present analyses adjusting for the randomization strata variables, while also providing unadjusted analyses.

All analyses were undertaken on an intention-to-treat basis. All available data for each subject were included regardless of whether the subject completed the MB-BP program or control condition. For example, of the 25 participants that had at least 2 absences from the MB-BP program, 13 (52%) took part in the 6-month follow-up assessments and were included in analyses. Of the 15 participants that withdrew/discontinued MB-BP, five took part in the 6-month follow-up assessments and were included in analyses. To account for missing data, multiple imputation was implemented using predicted mean matching of 14 baseline variables, imputing missing variables separately for each group, described elsewhere.^14,37^ Fifty imputations were generated, and GEE analysis was performed in each one.

All statistical analyses were conducted in SAS, version 9.4, except for multiple imputation that was carried out in R, version 4.2.2 using mice package.^38^

## Results

The study included 201 participants divided into a control group (n=100) and an MB-BP group (n=101). There were no substantial differences between MB-BP vs. control group in all baseline variables (**Table 1**). The average ages were 60.0 and 61.0 years for the control and MB-BP groups, respectively. Sex distribution was similar, with 58.0% of participants in the control group, and 59.4% in the MB-BP group identifying as female. The majority of participants were non-Hispanic White, comprising 83.0% of the control group and 79.2% of the MB-BP group. The primary outcome, CVH, had similar baseline scores according to the Lloyd-Jones et al method,^5^ of 64.0 in the control group and 65.6 in the MB-BP, which represents moderate levels of CVH.

**Table 1.** Baseline Characteristics of Study Sample by Treatment Group.

|  | Control Group (n=100) | MB-BP Group (n=101) |
| --- | --- | --- |
| **Age, years (mean, SD)** | 60.0 (12.3) | 61.0 (12.2) |
| **Sex (n, %)** |  |  |
| Male | 42 (42.0%) | 41 (40.6%) |
| Female | 58 (58.0%) | 60 (59.4%) |
| **Race/ethnicity (n, %)** |  |  |
| Asian | 2 (2.0%) | 3 (3.0%) |
| Black/African American | 2 (2.0%) | 7 (6.9%) |
| Hispanic | 5 (5.0%) | 3 (3.0%) |
| Native American | 0 (0.0%) | 3 (3.0%) |
| White | 83 (83.0%) | 80 (79.2%) |
| All other races/ethnicities | 5 (5.0%) | 5 (5.0%) |
| Don't know/refused | 3 (3.0%) | 0 (0.0%) |
| **Education (n, %)** |  |  |
| High School | 12 (12.2%) | 10 (10.2%) |
| Associate degree | 5 (5.1%) | 3 (3.1%) |
| College (4 year) | 28 (28.6%) | 34 (33.7%) |
| Graduate School | 43 (43.9%) | 41 (41.8%) |
| Other, n (%) | 10 (10.2%) | 10 (10.2%) |
| **Cardiovascular Health Components** |  |  |
| Cardiovascular Health Score, mean (SD) | 64.0 (10.2) | 65.6 (10.2) |
| Physical Activity, MET minutes/week, median (IQR) | 3321 (1603-6600) | 3397 (1855-6637) |
| DASH Diet Score, mean (SD) | 5.1 (1.5) | 5.3 (1.3) |
| Body Mass Index, kg/m<sup>2</sup>, mean (SD) | 29.5 (6.4) | 29.9 (6.6) |
| Sleep, hours per night, median (IQR) | 7.0 (6.0-8.0) | 6.5 (6.0-7.0) |
| Systolic Blood Pressure, mmHg, mean (SD) | 138.4 (13.7) | 138.6 (12.7) |
| Diastolic Blood Pressure, mmHg, mean (SD) | 82.6 (9.9) | 81.9 (8.4) |
| Cigarette Smoking, (n, %) |  |  |
| No | 39 (92.9%) | 33 (94.3%) |
| Yes | 3 (7.14%) | 0 (0%) |
| Don't know/refused | 0 (0%) | 2 (5.7%) |
| **Other Relevant Variables** |  |  |
| Sedentary Activity, sitting minutes/week, median (IQR) | 2220 (1560-3060) | 1980 (1440-3000) |
| Daily Alcohol Use, drinks/day, median (IQR) | 0.3 (0.1-1.0) | 0.4 (0.1-1.0) |
| Antihypertensive Medication Use (n, %) | 70 (70.1%) | 64 (64.0%) |
| Mindfulness (FFMQ) score | 133.4 (20.8) | 132.9 (19.2) |
| Perceived Stress Scale (PSS-14) score | 22.3 (8.6) | 23.6 (9.0) |

Of the 201 participants enrolled, 18 in the control group and 16 in the MB-BP group had no data collection at 6 months follow-up, resulting in an 82.6% follow-up rate. With the COVID pandemic affecting the 6-month follow-up in-person data collection for the last three cohorts, and CVH components not prioritized for data collection as they were not the preregistered primary outcomes (other than systolic blood pressure and diet), at least one CVH outcome variable was available at both baseline and the 6 month follow-up for 165 participants (82.1%; 81 MB-BP and 84 control); at least two CVH outcome variables were available at both baseline and the 6-month follow-up for 156 participants (77.6%; 78 MB-BP and 78 control). Four or more outcome variables were available for 131 participants (65.2%; 67 MB-BP and 64 control), and all six outcome variables were available for 109 participants (54.2%; 54 MB-BP and 55 control). Please see the CONSORT diagram in **Figure 2** for further details. Due to missing variables, findings prioritize reporting the multiple imputation results. Complete case analyses showed similar findings and are available in **Supplementary Table S2**.

**Figure 2.**
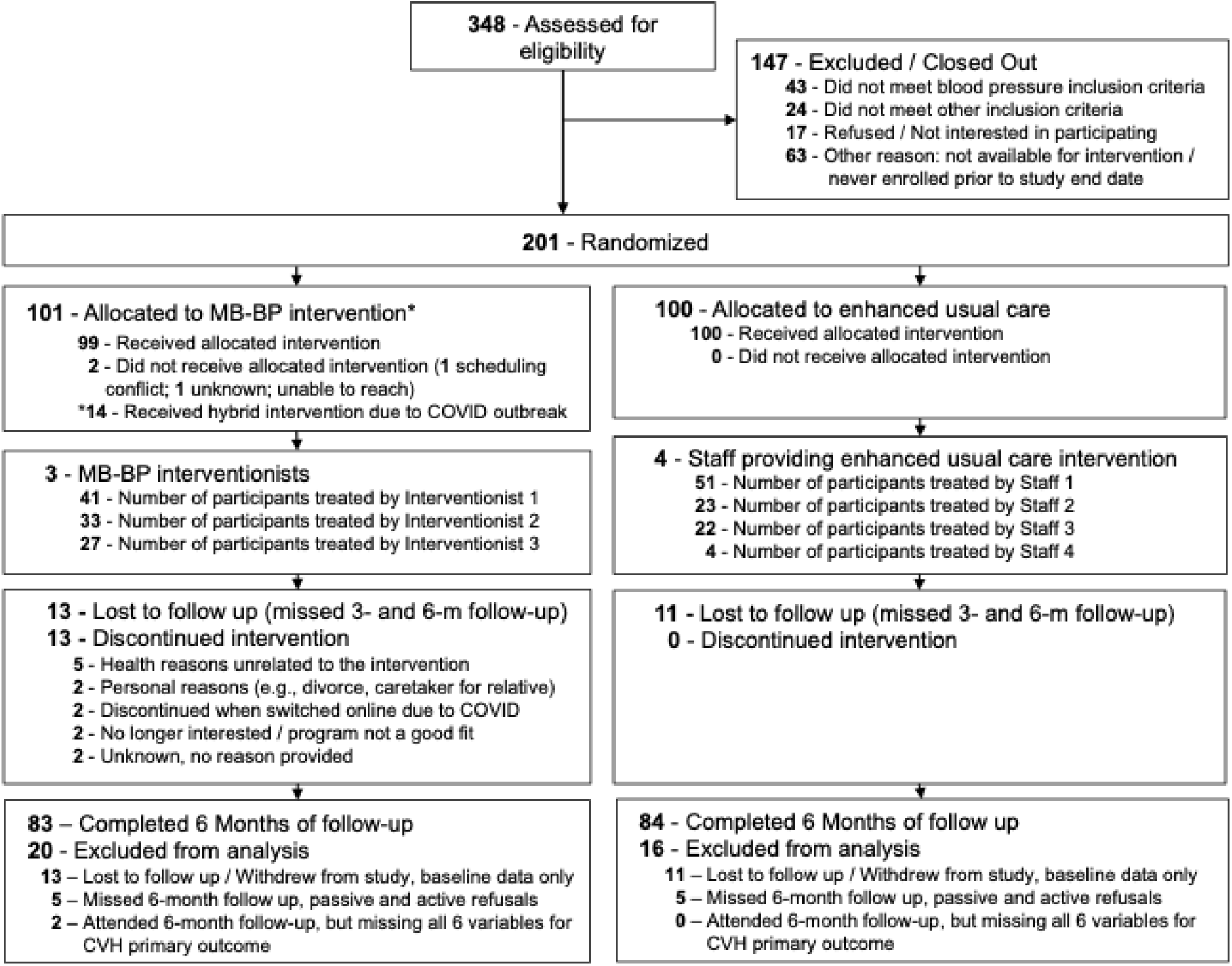
CONSORT diagram. Note that multiple imputation analyses were prioritized which resulted in all participants (n=201) included in primary analyses.

Adjusted analyses demonstrated that participants randomized to MB-BP had 0.144 greater standardized mean (95% CI: 0.023, 0.266; Cohen’s d=0.164) CVH scores by 6 months follow-up compared to control (**Figure 3**). MB-BP influenced all CVH components in healthier directions, with the exception of smoking which was close to null (0.05 cigarettes per day (95% CI: −0.05, 0.15) increase in MB-BP vs. control). Effects of MB-BP vs. control on CVH components were greatest for physical activity (47.9 MET min/week; 95% CI: −16.5, 112.3), sleep (0.34 hours/night; 95% CI: −0.10, 0.78), systolic blood pressure (−4.95 mmHg; 95% CI: −10.34, 0.44) and DASH diet score (0.27, 95% CI: −0.15, 0.69), as shown in **Table 2**. Unadjusted analyses showed similar findings (**Table 2**), as did complete case analyses (**Supplementary Table S2**). Effects of MB-BP vs. control on standardized means of all individual CVH variables are shown in **Supplementary Table S3**. Exploratory analyses utilizing the weighted categorical version of the CVH score showed similar directions of effect, while also demonstrating an expected lower sensitivity of the CVH variables to the intervention due to the variables being weighted categorical rather than continuous (**Supplementary Table S4**).

**Figure 3.**
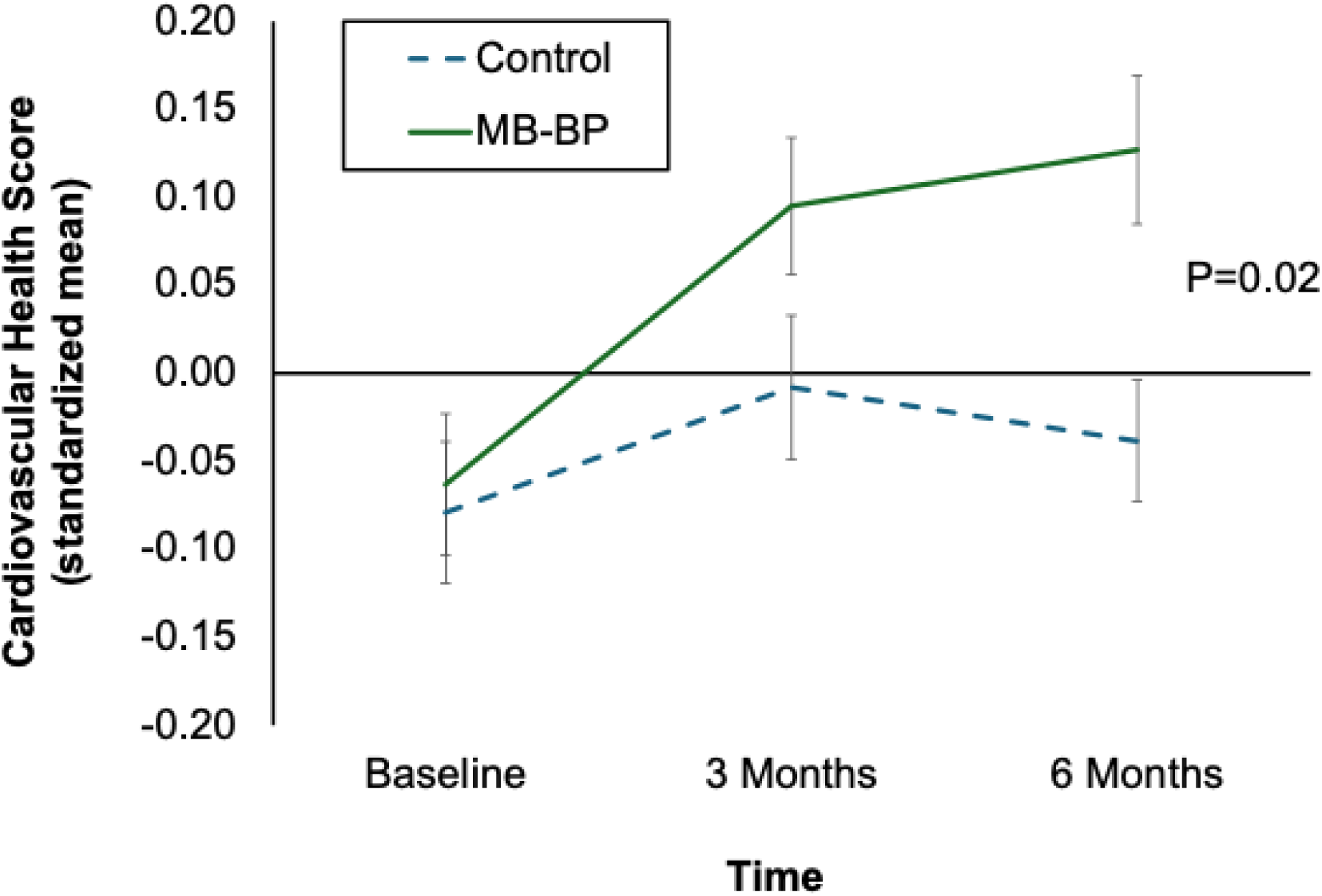
Impacts of MB-BP vs. control on cardiovascular health score. Error bars represent standard error of the mean.

**Table 2.** Regression analyses on effects of MB-BP vs. control on components of CVH. Outcome point estimates are continuous variables. Analyses were performed using multiple imputation. Adjusted analyses were controlled for baseline randomization strata (gender, baseline hypertension status).

| Outcome Variable | Timepoint |  |  |  |
| --- | --- | --- | --- | --- |
|  | 10 Weeks |  | 6 Months |  |
|  | Estimate (95% CI) | P | Estimate (95% CI) | P |
| **Unadjusted** |  |  |  |  |
| Physical Activity, MET min/wk | 25.1 (-33.6, 83.9) | 0.40 | 52.4 (-16.6, 121.3) | 0.14 |
| Cigarette Smoking, cigs/day | 0.05 (-0.05, 0.14) | 0.33 | 0.05 (-0.05, 0.15) | 0.34 |
| Sleep Duration, hr/night | 0.14 (-0.15, 0.62) | 0.23 | 0.33 (-0.04, 0.70) | 0.080 |
| Body Mass Index, kg/m<sup>2</sup> | 0.09 (-0.80, 0.98) | 0.84 | -0.28 (-1.29, 0.73) | 0.59 |
| Systolic Blood Pressure, mmHg | -2.6 (-7.6, 2.4) | 0.31 | -4.83 (-10.17, 0.51) | 0.077 |
| DASH Diet, score | 0.16 (-0.28, 0.60) | 0.47 | 0.28 (-0.10, 0.66) | 0.15 |
| **Adjusted** |  |  |  |  |
| Physical Activity, MET min/wk | 31.6 (-33.1, 96.4) | 0.34 | 47.9 (-16.5, 112.3) | 0.15 |
| Cigarette Smoking, cigs/day | 0.04 (-0.04, 0.13) | 0.33 | 0.05 (-0.05, 0.15) | 0.34 |
| Sleep Duration, hr/night | 0.25 (-0.08, 0.57) | 0.13 | 0.34 (-0.10, 0.78) | 0.13 |
| Body Mass Index, kg/m<sup>2</sup> | 0.09 (-0.80, 0.99) | 0.84 | -0.28 (-1.32, 0.75) | 0.59 |
| Systolic Blood Pressure, mmHg | -2.7 (-7.7, 2.2) | 0.28 | -4.95 (-10.34, 0.44) | 0.072 |
| DASH Diet, score | 0.17 (-0.25, 0.59) | 0.44 | 0.27 (-0.15, 0.69) | 0.20 |

Eight serious adverse events were observed during the 6-month follow-up period, with four occurring in the control group and four in the MB-BP group. Adverse events related to physical injuries were equally distributed between the study groups (n = 8 per group). No serious adverse events or physical injuries were determined to be related to study participation. Additional details on adverse events are reported elsewhere.^39^

## Discussion

This randomized controlled trial evaluated the impact of MB-BP on CVH and found significant improvements at 6 months follow-up among participants randomized to MB-BP compared to the control group. All CVH components showed positive trends, except smoking, which may be attributed to the small number of smokers (n=3) in the sample of 201 participants. These findings suggest that MB-BP is a promising intervention for CVH promotion.

### Comparison to Existing Literature

While self-reported mindfulness levels were found to be associated with cardiovascular health in a cross-sectional observational study,^40^ to our knowledge, no other mindfulness interventions targeting CVH as a comprehensive measure of both health behaviors (e.g., diet, physical activity, nicotine exposure, sleep) and health factors (e.g., BMI, blood pressure) have been published in the peer-reviewed literature. However, several studies have examined mindfulness effects on individual CVH components, as discussed below.

#### Blood Pressure

This study demonstrated reduced systolic blood pressure, consistent with previous findings.^13,41^ The effect was particularly evident in complete case analyses that did not adjust for baseline blood pressure stratification, likely in part because statistical adjustments for outcomes can reduce model precision and introduce bias when baseline levels are balanced between groups.^33–35^ Prior research supports mindfulness and meditation training as a means to lower blood pressure.^42^ However, further long-term follow-up studies with ambulatory blood pressure monitoring and diverse populations are needed.

#### Physical Activity

The modest improvement in physical activity (47 MET minutes per week) aligns with systematic reviews indicating that both dispositional mindfulness and mindfulness training support increased physical activity.^43,44^ MB-BP has also demonstrated significant effects on lowering sedentary behavior, reported elsewhere.^14,17^ Mindfulness may also enhance the affective experience of exercise, making physical activity more enjoyable.^45^ Interventions appear to be most effective when specifically designed for physical activity and when they target psychological factors influencing exercise behavior.^43^

#### Smoking

No significant effect on smoking cessation was observed, at least in part due to the small number of smokers (n=3) in the sample. Previous studies highlight mindfulness as a potential tool for smoking cessation, although findings remain mixed in systematic reviews.^46,47^

#### Sleep Duration

This study showed some evidence of improved sleep (20 minutes per night average improvement for MB-BP vs. control), although findings were not statistically significant (p = 0.196). Participants were not recruited based on low sleep duration, and baseline sleep levels (6.5–7.0 hours per night) were close to the recommended 7-9 hours per night for adults. Mindfulness-based interventions have shown effects in populations with inadequate sleep, such as those with insomnia.^48^ Mindfulness training may also help regulate sleep quality and duration, promoting wakefulness for those who oversleep (e.g., due to depression) and relaxation for those with insufficient sleep, in part by redirecting attention away from repetitive negative wakeful thoughts.^49^

#### Dietary Patterns

This study found modest non-significant evidence of improved diet quality, including greater DASH diet adherence. Effects were larger in participants of this study that had poor baseline diet quality, as reported elsewhere.^13,17^ While observational studies suggest that mindfulness is associated with healthier dietary patterns,^50,51^ few clinical trials have assessed its impact on dietary patterns such as DASH, Mediterranean diets, or caloric intake. Systematic reviews about the effects of mindfulness training on dietary patterns report mixed results.^44,52^ MB-BP may be particularly effective in supporting DASH diet adherence, as detailed in previous research.^13,17^

#### Adiposity

Minimal effects were observed on BMI (0.28 kg/m^2^ lowering by 6 months follow-up in MB-BP vs), which aligns with research indicating small-to-moderate effects from MBIs on weight-related outcomes.^53^ Since overweight/obesity or desire for weight loss were not an inclusion criterion, not all participants were seeking to lower adiposity. Additionally, among participants with overweight or obesity, not all prioritized weight loss, which may have limited intervention effects.

However, while BMI reductions in those who when through MB-BP have shown significant short-term effects, these effects have not persisted over the long term.^13,17^ Given that BMI has a substantial impact on blood pressure and that over 70% of Americans are overweight or obese, contemporary iterations of MB-BP have incorporated additional mindfulness-based practices focused on weight management. These include exploring participants’ relationships with evidence-based weight loss strategies, such as caloric restriction and GLP-1 inhibitor prescriptions.

We acknowledge ongoing advancements in recommendations regarding the use of BMI as a measure of adiposity at the individual level. Current guidelines suggest that excess adiposity should be confirmed either through direct measurement of body fat when available, or by including at least one additional anthropometric criterion (e.g., waist circumference, waist-to-hip ratio, or waist-to-height ratio), using validated methods and cutoff points appropriate for age, gender, and ethnicity.^54^

### Mechanisms

The MB-BP program likely improves CVH through enhanced self-regulation, including self-awareness, attention control, and emotion regulation. These mechanisms were originally hypothesized in the theoretical framework for MB-BP (**Figure 1**) and have been supported by both qualitative and quantitative findings.

#### Self-Regulation Pathways

In-depth interviews and focus groups revealed that self-awareness often serves as the first step in MB-BP-induced behavior change—for instance, recognizing stress or noticing the emotional impact of dietary choices.^55^ Participants then engaged in: (1) attention control (e.g., redirecting focus toward healthier behaviors) and (2) emotion regulation (e.g., using the STOP method—Stopping, Taking a breath, Observing thoughts, and Proceeding mindfully). These self-regulatory strategies enabled participants to adopt healthier behaviors, including improved diet, increased physical activity, and better stress management.^55^

#### Quantitative Findings on Self-Regulation

MB-BP participants demonstrated improvements in self-awareness (as measured by the Multidimensional Assessment of Interoceptive Awareness [MAIA]), emotion regulation (evidenced by reductions in depression symptoms and perceived stress), and attention control (as assessed using the Sustained Attention to Response Task [SART]).^13,17,56^ MB-BP also appears to have beneficial effects on multiple CVH-related components, including blood pressure, sedentary behavior, adherence to the DASH diet, and BMI.^13,17,57^

Although formal mediation analyses were not feasible due to sample size limitations, exploratory mediation analyses in this study sample, reported elsewhere, suggested that mindfulness and self-awareness each accounted for approximately one-third of the indirect effect of MB-BP on DASH diet adherence.^13^ Overall, by targeting multiple CVH risk factors, including diet, physical activity, and stress, MB-BP supports behavioral changes across CVH components, with the potential to reduce CVD risk and improve overall well-being.

### Clinical Implications

Findings from the MB-BP program demonstrated varying levels of clinical relevance across CVH outcomes. For systolic blood pressure, the observed reduction of 6 mmHg suggests a clinically meaningful effect. A systematic review and meta-analysis of 344,716 participants with elevated blood pressure in randomized controlled trials found that a 5 mmHg reduction in systolic blood pressure achieved through antihypertensive medication was associated with a 10% reduction in major cardiovascular events, such as myocardial infarction and stroke.^58^

Similarly, the observed improvement in the DASH score of 0.27 units should be interpreted in the context that participants were not recruited based on CVH parameters other than blood pressure. Consequently, many participants had healthy diets, body mass index (BMI), and physical activity levels at baseline. In a separate analysis of these data, restricting the sample to participants with initially unhealthy DASH diet scores revealed significant improvements—equivalent to increasing daily consumption by approximately one serving of fruits or vegetables.^13^

Physical activity improved by 48 MET-minutes per week, roughly translating to an additional 24 minutes of moderate pace walking per week (moderate pace walking has an MET score of about 3 METs compared to a person at rest that has 1 MET). A systematic review and meta-analysis found that an increase of 675 MET-minutes per week was associated with a 23% reduction in CVD mortality, suggesting that the more modest increase of 48 MET-minutes/week observed here may translate to approximately a 1.6% reduction in CVD mortality risk.^59^

The reduction in BMI of 0.28 kg/m² corresponds to an estimated 2-pound weight loss for an individual who is 5 feet 9 inches tall. A meta-analysis of over 900,000 participants found that among those with overweight or obesity, each 1 kg/m² increase in BMI was associated with a 6% increase in all-cause mortality and an 8% increase in vascular mortality. Thus, a 0.28 kg/m² reduction may represent approximately a 1–2% decrease in all-cause mortality risk and a 2–3% reduction in vascular mortality risk.^60^

The increase in sleep duration of 0.34 hours corresponds to about 20 additional minutes of sleep per night. Given that the average baseline sleep duration in the study was 6.5 to 7.0 hours—slightly below the recommended 7 to 9 hours to minimize CVD risk—this improvement is potentially meaningful.^5,61^

Taken together, these modest improvements across multiple drivers of CVH and CVD risk suggest a potential cumulative impact on reducing CVD risk and enhancing overall well-being. Additional analyses from this dataset reported elsewhere have demonstrated significant improvements in other markers of cardiovascular and whole-person health, including reductions in sedentary behavior and depressive symptoms.^14,56^

### Study Limitations

This study includes several limitations. Firstly, the measure of CVH did not include glucose regulation or lipids, due to the study not collecting blood samples. Including these components in future studies would provide a more comprehensive understanding of how mindfulness training affects cardiovascular health.^5^ Secondly, the limited diversity of the sample, with a predominantly non-Hispanic White and well-educated population, restricts generalizability, highlighting the need to include more diverse populations in future research using evidence-based strategies.^62–64^ The program shows promise as a culturally adaptable approach to delivering blood pressure–specific health education, potentially more targeted than standard MBSR.^39,64,65^ Current work by our team focuses on adapting and evaluating the program in more diverse populations, cultures, and languages – including Native American populations and those living with HIV/AIDS. Thirdly, the 6-month follow-up period is too short to assess the long-term sustainability of the program’s effects, necessitating longer-term studies to evaluate whether improvements in CVH persist over time. However, in a prior clinical trial of MB-BP, effects on systolic blood pressure were shown to hold through at least 2 years follow-up.^41^ Fourthly, while standard recommendations for measuring CVH by the Life’s Essential 8 construct recommend utilizing a scoring system that provides points to categories of each construct (described in the Outcomes subsection in the Methods section),^5^ this study showed evidence that continuous variables were more responsive to the intervention than the Life’s Essential 8 categorically-derived point scores, which would be expected as continuous variables typically increase statistical power and sensitivity to interventions compared to categorically-derived scores. Future measurement recommendations for CVH may want to consider the use of continuous variables rather than categorically-derived scores in clinical trials, such as through standardized means, to offer greater sensitivity of the Life’s Essential 8 score to interventions.

### Conclusions

Findings indicated that a structured mindfulness intervention, specifically MB-BP, was effective in supporting CVH, with evidence that most CVH components (e.g., systolic blood pressure, DASH diet, sleep duration, body mass index) modestly shifted in healthier directions. If these findings are replicated with longer term follow up, and evaluated in diverse populations and health care settings, this could indicate that MB-BP has the potential to foster CVH, and contribute to lowering risk for cardiovascular disease, the primary cause of death world-wide.

## Data Availability

Deidentified data for the primary outcome will be provided on the Open Science Framework website upon publication.

https://osf.io/86ucd/

